# Pharmacokinetics of favipiravir in adults with mild COVID-19 in Thailand

**DOI:** 10.1101/2022.03.09.22271220

**Authors:** Weerawat Manosuthi, Ing-orn Prasanchaimontri, Suvimol Niyomnaitham, Rujipas Sirijatuphat, Lantharita Charoenpong, Katherine Copeland, Tim R. Cressey, Phongpan Mokmued, Kulkanya Chokephaibulkit

**Affiliations:** Bamrasnaradura Infectious Diseases Institute, Department of Disease Control, Ministry of Public Health, Nonthaburi, Thailand; Bureau of Drug and Narcotic, Department of Medical Sciences, Ministry of Public Health, Nonthaburi, Thailand; Department of Pharmacology, Faculty of Medicine Siriraj Hospital, Mahidol University, Thailand; Siriraj Institute of Clinical Research (SICRES), Mahidol University, Thailand; Department of Medicine, Faculty of Medicine Siriraj Hospital, Mahidol University, Thailand; Mahidol University International College, Salaya, Nakhon Pathom, Thailand; AMS-IRD collaboration, Department of Medical Technology, Faculty of Associated Medical Sciences, Chiang Mai University, Thailand; Department of Molecular & Clinical Pharmacology, University of Liverpool, UK; Division of Genomics Medicine and Innovation Support, Department of Medical Sciences, Ministry of Public Health, Nonthaburi, Thailand; Department of Paediatrics, Faculty of Medicine Siriraj Hospital, Mahidol University, Thailand

**Keywords:** Favipiravir, COVID-19, Thai adults, pharmacokinetic

## Abstract

We assessed the pharmacokinetics of favipiravir (FPV) in adults with symptomatic SARS-CoV-2 infection without pneumonia in Thailand. FPV dosing was 1800 mg twice-daily on day 1, then 800 mg twice-daily for 14 days. Eight subjects (7 female), median (range) age 39 (19-53) years and BMI 27.9 (18.0-33.6) were included. Inter-subject variability was high but all achieved minimum plasma concentrations (C_min_) above EC_50_ (9.7 mg/L). FPV was well tolerated; 1 subject stopped prematurely due to rash.

## Introduction

Favipiravir (FPV) is a pyrazine carboxamide derivative that has recently been repurposed to treat mild-to-moderate cases of COVID-19 (1,2). Preliminary studies have demonstrated its capacity to competitively inhibit the replication of SARS-CoV-2 *in-vitro* (3-5) and possibly control viral progression, promote viral clearance, as well as improve clinical outcomes *in-vivo* (5, 6).

FPV is a prodrug (T-705) that is intracellularly metabolised through ribosylation into its active form (T-705-RTP). Viral RNA-dependant RNA polymerase (RdRp) recognises this active form as a purine-base analogue and incorporates it into nascent viral RNA whereby it exerts its antiviral activity (1, 2, 5-7). FPV is primarily metabolised by hepatic aldehyde oxidase, and partially by xanthine oxidase, to a hydroxylated inactive metabolite (T-705M1) that is excreted in the urine (4, 7, 8).

The recommended FPV dosing regimen for the treatment of COVID-19 in Thailand is 1800 mg twice-daily for the first day, and then 800 mg twice-daily for a total duration of 5-14 days (5). While numerous studies have explored the clinical effects of such dosing regimens, few studies have assessed its pharmacokinetics, and no studies have been performed in the Thai population (9, 10). It is important to characterise the impact of regional and ethnic difference on the pharmacokinetic profile to ensure optimised dosing for the viral variants circulating in the region (9). Our aim was to investigate the pharmacokinetics of FPV in mild cases of COVID-19 without pneumonia within a Thai population.

## Methods

Here we report the results of a pharmacokinetic sub-study of FPV nested within a prospective randomized clinical trial of adults with symptomatic SARS-CoV-2 infection without pneumonia. Enrolled subjects were randomised to receive supportive symptomatic care with or without FPV. The FPV dosing regimen was 1800 mg twice-daily on the first day, and then 800 mg twice-daily for 5-14 days or until viral clearance. Subjects receiving other medication with reported antiviral activity against SARS-CoV-2 were excluded. Pharmacokinetic assessments were proposed to subjects receiving FPV. This study was approved by the Siriraj Institutional Review Board (approval no. Si 434/2020) and registered at Thaiclinicaltrials.org (No. TCTR20200514001).

### Pharmacokinetic Assessments

Blood samples were drawn pre-dose, and 1-hour after FPV administration on days 1, 2, 3, 4, 5, 7, and 14. Plasma proteins were precipitated with ethanol (1:4) and the supernatant stored at -20°C until analyses. T-705 and T-705M1 concentrations were quantified using a validated high performance liquid chromatography-UV assay, with a calibration range of 0.1 to 50 mg/L.

## Results

Eight patients (7 females) were included, median (range) age was 39 (19-53) years, BMI was 27.9 (18.0-33.6), 3 patients (37.5%) had underlying diseases, and none developed pneumonia or experienced disease progression. Subjects No. 1 and 3-5 were treated with FPV for 14 days, while subjects No. 6-8 were treated for 7 days. Subject No. 2 dropped out on day 4 of treatment due to an erythematous skin rash. Individual plasma concentrations versus time curves for T-705 and T-705M1 after the first dose of FPV are shown in Figure 1. Initial pre-dose T-705 concentrations for subjects No. 4 and 5 indicated they had received a dose of FPV before PK assessment.

**Figure 1.**
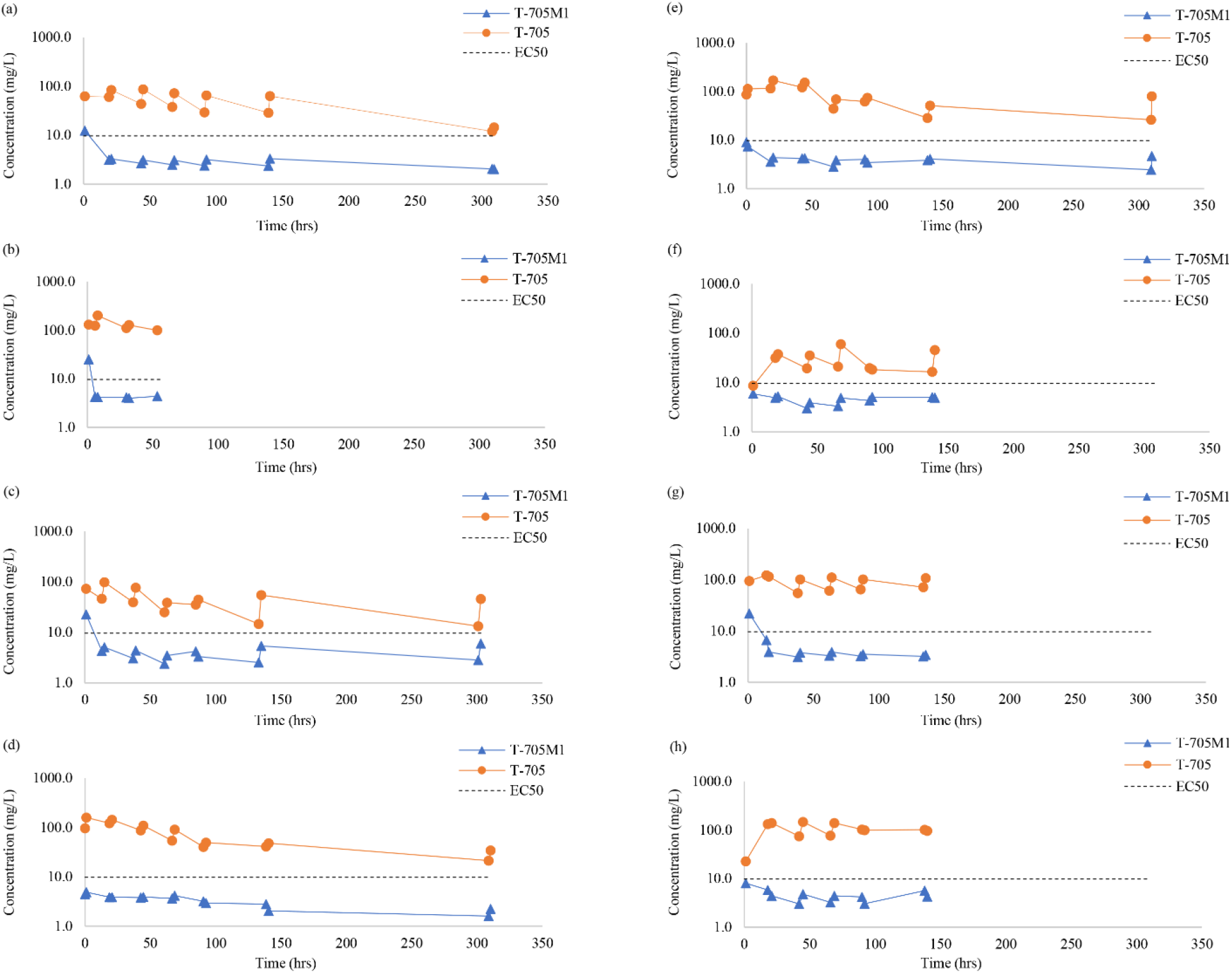
Individual plasma concentration of T-705 and T-705M1 for (a) Subject No. 1, female with BMI of 30.1 within age range of 30-35 years, (b) Subject No. 2, female with BMI of 18.3 within age range of 35-40 years, (c) Subject No. 3, female with BMI of 33.6 within age range of 40-45 years, (d) Subject No. 4, female with BMI of 30 within age range of 25-30 years, (e) Subject No. 5, female with BMI of 27.5 within age range of 45-50 years, (f) Subject No. 6, male with BMI of 26.9 within age range of 50-55 years, (g) Subject No. 7, female with BMI of 28.2 within age range of 35-40 years, and (h) Subject No. 8, female with BMI of 18 within age range of 15-20 years. The dash line represents an EC_50_ of 9.7 mg/L.

Minimum plasma concentration levels (C_min_) of T-705 were above the reported EC_50_ for SARS-COV-2 (9.7 mg/L) for all subjects, except for subject No. 6 after their first dose (8.6 mg/L). The C_min_ and C_max_ of T-705 ranged from 12.1-132.2 mg/L and 14.5-201.2 mg/L, respectively, and was found to decreased overtime. C_min_ and C_max_ of T-705M1 ranged from 1.6-6.7 mg/L and 2.0-24.8 mg/L, respectively, but remained relatively stable overtime. Inter-subject variability was high, with a coefficient of variation (CV) of 37-74% for T-705 and 12-62% for T-705M1.

## Discussion

We evaluated FPV plasma concentrations in adults with mild COVID-19 that did not require oxygen supplementation. FPV plasma concentrations with the currently recommended FPV dosing regimen in Thailand [1800 mg twice-daily (9 tablets per dose) on day 1, followed by 800 mg twice-daily (4 tablets per dose) until the end of treatment] provided concentrations above the reported EC_50_.

Favipiravir loading doses studied have ranged from 400-6000 mg, split into 2-3 doses per day (11). A 1600 mg twice-daily dose on day 1, followed by 600 mg twice-daily on days 2-5 achieve trough concentrations between 20-60 mg/L after 12 hours (12). The large loading dose of FPV used in our study was reported to rapidly reach minimum target concentrations of 40-80 mg/L, and the large difference between its EC_50_ of 61.88 μM (9.72 mg/L) and half-cytotoxic concentration (CC_50_) of >400 μM (62.83 mg/L) provides a wide therapeutic range (3,5,11).

Some controversy remains regarding the EC_50_ of FPV for SARS-CoV-2. Some studies have reported similar values to our study (9,12); while others failed to identify inhibition of SARS-CoV-2 below 100 μM (15.71 mg/L) *in-vitro* (15), or found only 50% virus inhibition at this concentration (16). All of the adults in our study achieved T-705 concentrations above our target EC_50_ of 9.7 mg/L. The high interpatient variability observed is likely a result of the multiple intrinsic and external factors that can impact the pharmacokinetic profile of FPV (9). For example, the potential impact of pharmacogenomics on FPV clearance in different populations is highlighted by the significantly higher FPV plasma concentrations in Japanese populations compared to patients in the United States (7), and decreased exposures in African and American patients compared to Chinese patients (18). While the T-705 plasma concentration varied between patients, the overall concentrations observed in our study remained within target ranges. Favipiravir was well tolerated, expect for one patient who experienced a cutaneous rash and discontinued treatment. Of note, this participant had relatively high FPV concentrations (99.7-201.2 mg/L) and low BMI (18.3). None of the patients’ clinical outcomes deteriorated during FPV treatment but a larger population of mild COVID-19 cases is needed to confirm this observation.

A limitation of this study was the ability to associate clinical or virologic outcomes with FPV drug exposure. Overall, this study provides novel data on the pharmacokinetics of FPV and its metabolite in adults with symptomatic SARS-CoV-2 infection without pneumonia and can help to guide dosing recommendations.

## Acknowledgements

We would like to thank the National Centre for Global Health and Medicine, Japan and FUJIFILM Toyama Chemical Co., Ltd. We are also thankful for the support from the National Institute of Health, Thailand.

## Data availability statement

Data are available upon request.

## Transparency declarations

None to declare

